# Measuring T-Cell Response to SARS-CoV-2 Using Elecsys IGRA SARS-CoV-2 Assay

**DOI:** 10.1101/2025.05.19.25327215

**Authors:** Markus Eckl, Chih-Yeh Chen, Ulrike Schulte-Spechtel, Qian Feng

**Author notes:** **Corresponding author:** Qian Feng, Forrenstrasse 2, 6343 Rotkreuz, Switzerland.

## Abstract

**Purpose:** This study aims to assess T-cell response in individuals with a previous severe acute respiratory syndrome coronavirus 2 (SARS-CoV-2) infection or coronavirus disease 2019 (COVID-19) vaccination using the Elecsys^®^ IGRA SARS-CoV-2 assay that is designed to detect broad-spectrum T-cell response. Preliminary data show that SARS-CoV-2 can elicit a robust and durable T-cell response, which, in conjunction with antibody response, may contribute to infection control. Furthermore, studies have shown that T-cell response is not substantially affected by emerging variants. The Elecsys IGRA SARS-CoV-2 assay combines *in vitro* T-cell stimulation and an automated electrochemiluminescence immunoassay for interferon-γ measurement to qualitatively detect T-cell-mediated response to SARS-CoV-2 in human whole blood.

**Methods:** Clinical sensitivity of the Elecsys IGRA SARS-CoV-2 assay in convalescent and vaccinated adults was evaluated in a prospective clinical study including two cohorts in Germany, while specificity was determined in participants who were unvaccinated, without prior knowledge of a SARS-CoV-2 infection, and non-reactive to the Elecsys Anti-SARS-CoV-2 and Elecsys Anti-SARS-CoV-2 S serology assays.

**Results:** The Elecsys IGRA SARS-CoV-2 assay showed high sensitivity and specificity in detecting SARS-CoV-2-elicited T-cell response. The Elecsys IGRA SARS-CoV-2 assay was found to reliably detect T-cell response upon natural infections or vaccinations, with sensitivity exceeding 99%, which was maintained up to 307 days following the last dose of the vaccine and up to 736 days post-symptom onset.

**Conclusion:** Measuring T-cell response in convalescent and vaccinated individuals is feasible and can be meaningful in providing a more complete picture of the immune status of individuals in conjunction with serology assays.

## Introduction

An outbreak of a novel coronavirus has spread worldwide, which led the World Health Organization (WHO) to declare a public health emergency of international concern in early 2020 [1, 2]. Coronavirus disease 2019 (COVID-19) is caused by an infection with severe acute respiratory syndrome coronavirus 2 (SARS-CoV-2) [3] and, as of January 2025, there have been more than 760 million confirmed cases of COVID-19, including 6.9 million deaths reported to WHO [4]. The clinical spectrum of SARS-CoV-2 infection is wide, ranging from an asymptomatic infection to severe pneumonia with acute respiratory distress syndrome and death [5]. Early diagnosis can contribute to clinical management and outbreak control. Diagnostic testing involves detecting the virus itself or detecting the human immune response to viral infection [6].

Commonly used diagnostic methods for SARS-CoV-2 include real-time (RT) polymerase chain reaction (PCR), which has become the gold standard; rapid (antigen) detection tests; viral culture; and serology assays [6]. The evaluation of prior SARS-CoV-2 infection and existing immunity, as well as epidemiologic studies of COVID-19, are currently reliant upon serologic testing of immunoglobin (Ig)M, IgG, and/or IgA antibody isotypes [7–9]. Since viral load in infected individuals generally decreases and cannot be reliably detected after the first week(s) post-symptom onset [10], serologic antibody testing identifies a wider swathe of exposures than molecular diagnostics [7]. Binding antibody assays are readily available and easy to perform, and hence used more often. On the other hand, neutralizing assays can assess the critical step of infection entry into the cell and provide functional information [11, 12]. These serology assays measure different immunoglobulin classes (IgM, IgA, IgG, or total Ig), affinities, and antigenic targets (epitopes) with various detection methods and assay formats.

However, the immune response is multifaceted, and it includes not only antibodies, but also cell-mediated responses. The immunologic basis for the human body’s heterogeneous response to infection is mediated by a range of soluble factors and immune cells, including T lymphocytes. T cells are increasingly recognized for their role in SARS-CoV-2 infection and immunity, and measuring the presence of T-cell responses can provide information that is complementary to serology [8, 13]. Poly-functional, largely interferon-gamma (IFN-γ)–secreting CD4^+^ T cells bestow SARS-CoV-2–specific T-cell immunity that is maintained throughout convalescence [14]. Infection and vaccination-elicited SARS-CoV-2–reactive CD4^+^ T cells appear to be broadly T helper 1 (Th1)-like, secreting IFN-γ, tumor necrosis factor-alpha (TNF-α), and interleukin-2 (IL-2) cytokines, while CD8^+^ T cells from COVID-19 patients have an activated phenotype and show cytotoxic potential. Cellular immunity supports host defenses against viruses in addition to humoral immunity [8, 15]. SARS-CoV-2–induced T cell responses are broadly detectable across disease stages and severities, age groups, and vaccine types [14, 16–18].

A recent study demonstrated that 75% of asymptomatic COVID patients had a detectable T-cell response at 15 months after infection, indicating the persistence of the T-cell signal [19]. Other studies have demonstrated a T-cell response at 9–13 months post infection for critically ill patients [20] and 6–8 months post infection for mild or moderate disease [21, 22]. A T-cell response was also observed at 6 months post vaccination or infection in vaccinated healthcare workers [23] and in patients undergoing immunomodulatory therapy [24]. Notably, SARS-CoV-2–reactive T cells have been detected in 20–50% of individuals without known exposure to the virus [25–27], and T-cell responses against SARS-CoV-2 have also been detected in recovered COVID-19 patients with no detectable antibodies [8]. Preliminary data thus show that SARS-CoV-2 can elicit robust and durable T-cell responses, which, in conjunction with antibody responses, may contribute to infection control.

Furthermore, T-cell responses are likely robust against mutations in emerging variants [28]. T-cell responses are focused not only on spike (S), but also on membrane (M), nucleocapsid (N), and non-structural proteins (NSPs) [27]. Ninety-three per cent and 97% of CD4 and CD8 epitopes are 100% conserved across variants [29]. Multiple studies have shown that 70–85% of the CD4^+^ and CD8^+^ T-cell response to the SARS-CoV-2 Omicron variant (B.1.1.529) is maintained across study groups who received different vaccines [6, 29]. T-cell responses to early SARS-CoV-2 variants were preserved across vaccine platforms, but memory B cells and neutralizing antibodies decreased significantly [29]. Similar findings were also noted in a cohort of 5- to 12-year-olds vaccinated with BNT162b2; this suggests that booster vaccinations do not confer additional immunological protection on healthy children [30].

T-cell immunity has even been observed in immunocompromised patients up to 6 months post infection [31]. Humoral and cellular responses in mRNA-vaccinated individuals were well correlated at 3 months post vaccination for healthy controls, people living with human immunodeficiency virus and patients with primary immunodeficiencies [32, 33].

SARS-CoV-2–reactive T-cell responses have been measured after vaccination in multiple sclerosis patients treated with ocrelizumab [34], in patients with B-cell–depleted lymphoma after chimeric antigen receptor T-cell therapy [35, 36], in patients undergoing immune-modifying therapies [24], and also in liver and heart transplant recipients [37]. Almost all healthy controls, 89% of individuals with inborn errors of immunity, and 76% with secondary immunodeficiency had an IFN-γ level above the validated reference range after T-cell stimulation following primary vaccination [38]. SARS-CoV-2–reactive T-cell responses can thus be measured in immunocompetent and in immunocompromised individuals.

Also of interest, individuals diagnosed with COVID-19 have been found to generate CD8+ and CD4+ T-cell responses [27, 39] associated with protection from infection and less severe disease, while peripheral T-cell lymphopenia is typically detected in symptomatic adult COVID-19 cases [13, 16]. T cells appear to be depleted in the peripheral blood of adult COVID-19 patients, and the magnitude of this reduction is positively correlated with disease severity, whereas peripheral T-cell counts are largely preserved in asymptomatic patients and children [13]. Notably, in COVID-19 infected patients with hematologic cancer, patients with a higher CD8 T cell count had improved survival, including those who received anti-CD20 therapy [40]. Rapid induction and magnitude of humoral responses is associated with greater disease severity, but early induction of IFN-γ–secreting SARS-CoV-2– specific T cells is associated with mild disease and accelerated viral clearance [41]. In patients with chronic kidney disease, lower levels of antibody titer and T-cell response were associated with significantly increased probability of subsequent infection [42]. Several studies show that CD3^+^, CD4^+^, and CD8^+^ T-lymphocyte counts may be inversely related to the severity of COVID-19 and monitoring T-cell response could help assess the potential for patients to become critically ill [13, 43].

The focus of SARS-CoV-2 diagnostic testing has begun to coalesce around immunological correlates of protection from disease based on humoral and cellular immunity [44]. Available technologies to detect T cells or immune cells include direct detection of surface markers, soluble marker detection, and functional release assays. The first category of tests may resort to detection of the surface marker at the protein level (e.g., cytometry by time-of-flight), at the DNA level (e.g., T-cell receptor sequencing), or use surface marker-specific antibodies conjugated to a magnetic or fluorescent detector [4, 45, 46]. Direct, cross-sectional soluble marker detection includes fluorescence-activated cell sorting and enzyme-linked immunoassays. Functional release assays, such as IFN-γ release assays (IGRAs) involve a stimulation step prior to measurement of a functional response, either by detecting a soluble cytokine in bulk or by measuring the number of cells producing the cytokine.

IFN-γ is a crucial cytokine for T-cell mediated immune response [47, 48]. When pathogen-derived molecules stimulate local IFN-γ production, IFN-γ augments the immune system response through an amplification loop to increase immune system sensitivity [49]. There is accumulating evidence that indicates that using IGRA to assess T-cell immunity may be a reliable test to be included in COVID-19 monitoring protocols, particularly after vaccination [30, 45, 50, 51]. The clinical utility of IGRA benefits from both the relative simplicity of the test compared to other alternatives (e.g., ELISPOT, flow cytometry cytokine secretion analysis, MHC tetramers, or activation-induced marker assays) and the familiarity of some laboratories with this technique, which is already employed for the diagnosis of latent tuberculosis infection [50, 52].

We aimed to determine the clinical performance of the Elecsys^®^ IGRA SARS-CoV-2 assay in detecting broad-spectrum T-cell response via IFN-γ measurement in whole blood samples from individuals with or without prior SARS-CoV-2 infection or COVID-19 vaccination. This test may provide valuable insights into the SARS-CoV-2 immune response and thus support both clinical management and population surveillance.

## Methods

The Elecsys IGRA SARS-CoV-2 assay combines *in vitro* T-cell stimulation and an automated electrochemiluminescence immunoassay (ECLIA) for IFN-γ measurement to qualitatively detect T-cell–mediated response to SARS-CoV-2 in human whole blood.

The Elecsys IGRA SARS-CoV-2 assay requires two steps as follows:

Step 1: *ex-vivo/in vitro* stimulation of patient T cells with pathogen-specific peptides to induce production of activation markers, including IFN-γ, if reactive T cells are present, indicating previous exposure of the patient to the antigen. This phase takes place in the Cobas^®^ IGRA SARS-CoV-2 tubes containing the patient’s whole blood specimen.

Step 2: IFN-γ read out in the plasma extracted from the whole blood mentioned in step 1 after incubation. This can be done either directly out of the Cobas IGRA SARS-CoV-2 tubes after centrifugation or after transfer of the separated plasma into a secondary tube.

The clinical sensitivity of the Elecsys IGRA SARS-CoV-2 assay in convalescent and vaccinated adults was evaluated in a prospective, all-comers clinical study including two cohorts in Germany from March to December 2022. Enrolled participants had either a positive RT-PCR diagnosis for SARS-CoV-2 infection in the past or had been vaccinated against COVID-19, but not both.

Cohort 1 (convalescents) consisted of 100 participants aged 21–65 years who had previously received at least one positive SARS-CoV-2 test result using a CE-marked RT-PCR method, but who had not been vaccinated at the time of this study. Cohort 2 (vaccinees) consisted of 296 participants aged 18–83 years who were fully vaccinated against COVID-19, including boosters, but who had never received a positive SARS-CoV-2 test result at the time of this study. Assay performance was also stratified based on demographic information (age and sex). Specificity of the Elecsys IGRA SARS-CoV-2 assay was determined in 78 participants who were unvaccinated, without prior knowledge of a SARS-CoV-2 infection, and non-reactive Elecsys Anti-SARS-CoV-2 and Elecsys Anti-SARS-CoV-2 S assays. These patients were recruited via blood donation services.

## Results

### Sensitivity

The Elecsys IGRA SARS-CoV-2 assay was found to reliably detect T-cell response upon natural infections or vaccinations, with sensitivities exceeding 99%. See **Table 1** for further details.

**Table 1.** Clinical sensitivity of the IGRA SARS-CoV-2 assay. IGRA, interferon-gamma release assay; SARS-CoV-2, severe acute respiratory syndrome coronavirus 2.

| Cohort | N | Reactive | Non-reactive | Indeterminate<sup>a</sup> | Invalid<sup>b</sup> | Clinical sensitivity,<br>% (95% CI)<sup>c</sup> |
| --- | --- | --- | --- | --- | --- | --- |
| Convalescent | 100 | 96 | 0 | 3 | 1 | 100 (96.2–100) |
| Vaccinees | 296 | 288 | 1 | 6 | 1 | 99.7 (98.1–100) |

<sup>a</sup>High baseline IFN-γ in negative control
<sup>b</sup>Positive control below threshold, indicative of insufficient number of fit T cells
<sup>c</sup>Indeterminate and invalid samples were excluded from the sensitivity calculations as they were unevaluable
95% CI determined using Clopper–Pearson exact method
CI, confidence interval; IFN-γ , interferon-gamma; IGRA, interferon-gamma release assay; SARS-CoV-2, severe acute respiratory syndrome coronavirus 2

T-cell response to SARS-CoV-2 was detected up to 736 days post-symptom onset (DPSO) and up to 307 days post-last vaccine dose (DPLastVax), respectively. The distribution over DPSO and DPLastVax of reactive cases closely paralleled the distribution of overall enrolled cases (**Fig. 1** and **Fig. 2**), suggesting that the time span since the last known exposure did not have a strong adverse effect on assay sensitivity in this study.

**Fig. 1.**
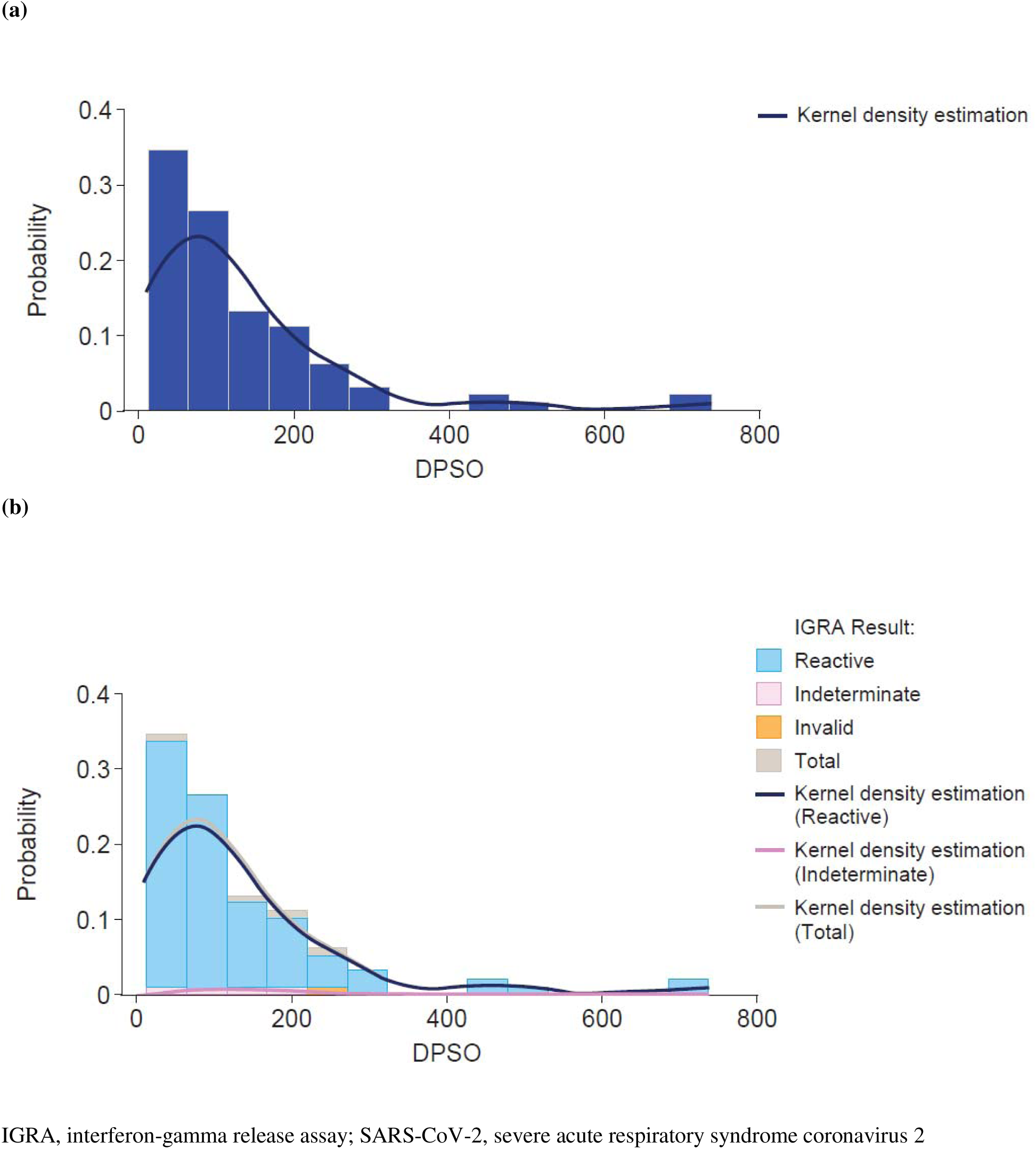
Distribution of days post-symptom onset in Cohort 1 (convalescents). **(a)** Overall distribution of all enrolled participants in Cohort 1. **(b)** Distribution stratified by Elecsys IGRA SARS-CoV-2 assay results. IGRA, interferon-gamma release assay; SARS-CoV-2, severe acute respiratory syndrome coronavirus 2

**Fig. 2.**
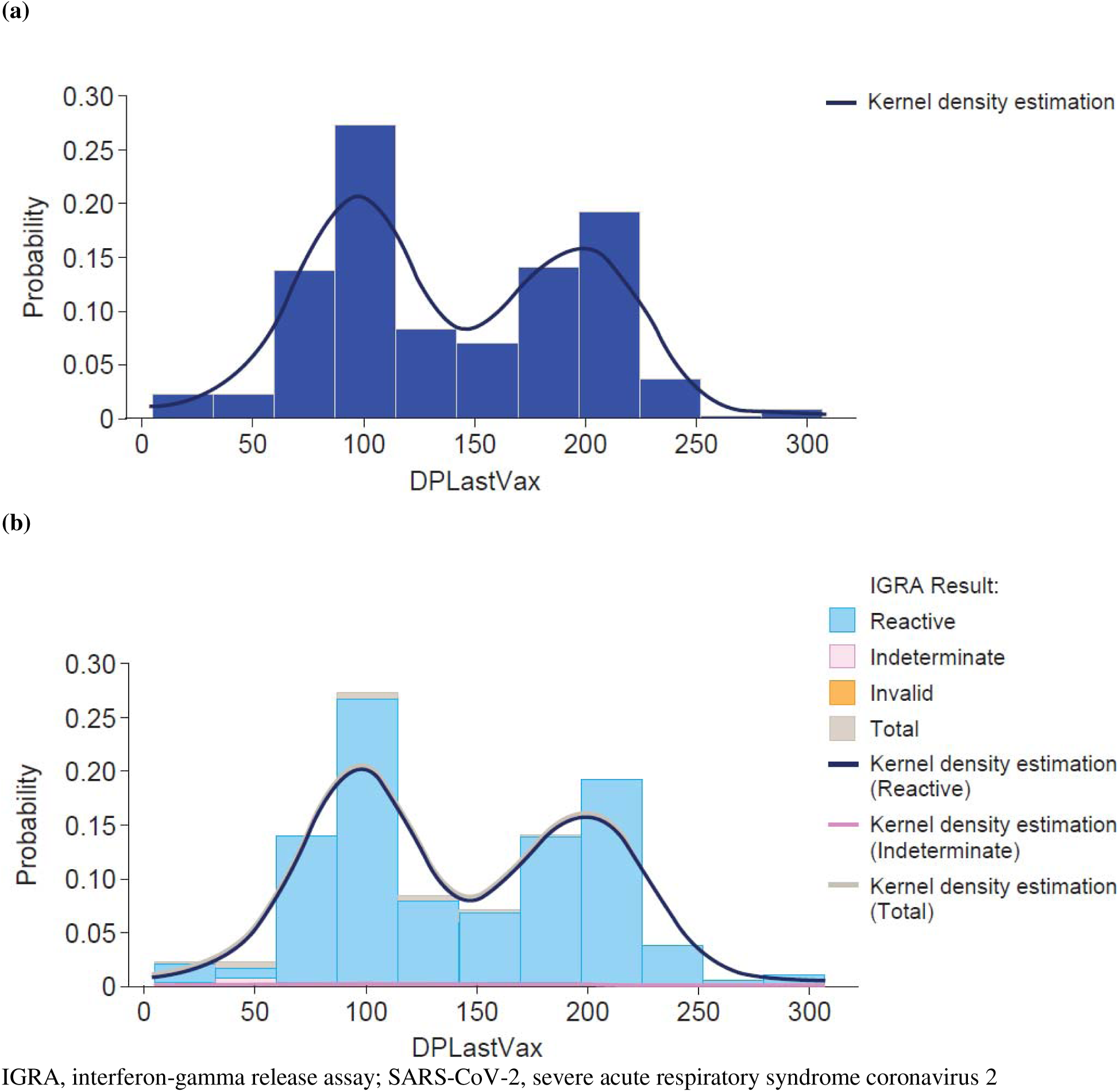
Distribution of days post-vaccination in Cohort 2 (vaccinees). **(a)** Overall distribution of all enrolled participants in Cohort 2. **(b)** Distribution stratified by Elecsys IGRA SARS-CoV-2 assay results. IGRA, interferon-gamma release assay; SARS-CoV-2, severe acute respiratory syndrome coronavirus 2

Furthermore, antibody responses in the cohorts of convalescents and vaccinees were also assessed using Elecsys Anti-SARS-CoV-2 and Elecsys Anti-SARS-CoV-2 S assays. Three participants in the convalescent cohort had discrepant Elecsys Anti-SARS-CoV-2 and Elecsys IGRA SARS-CoV-2 assay results. Among these, 3 participants were reactive for IGRA but non-reactive for Elecsys Anti-SARS-CoV-2 assay. In addition, 3 participants were indeterminate for IGRA and reactive for Elecsys Anti-SARS-CoV-2 assay (**Table 2**).

**Table 2.** IGRA test results compared to Anti-SARS-CoV-2 assay results in Cohort 1 (convalescents). Anti-SARS-CoV-2 assay detects antibodies targeting the SARS-CoV-2 nucleocapsid protein. IGRA, interferon-gamma release assay; SARS-CoV-2, severe acute respiratory syndrome coronavirus 2.

| IGRA result | Anti-SARS-CoV-2 reactive | Anti-SARS-CoV-2 non-reactive |
| --- | --- | --- |
| Reactive | 93 | 3 |
| Non-reactive | 0 | 0 |
| Indeterminate | 3 | 0 |
| Invalid | 1 | 0 |

All vaccinees had reactive results on the Elecsys Anti-SARS-CoV-2 S assay, regardless of their Elecsys IGRA SARS-CoV-2 assay results (Table 3). Stratification by baseline demographic information did not alter assay performance. The comparison of qualitative Elecsys IGRA SARS-CoV-2 assay results and quantitative Elecsys Anti-SARS-CoV-2 S assay results in the cohorts of convalescents and vaccinees is depicted in **Fig. 3**.

**Table 3.**
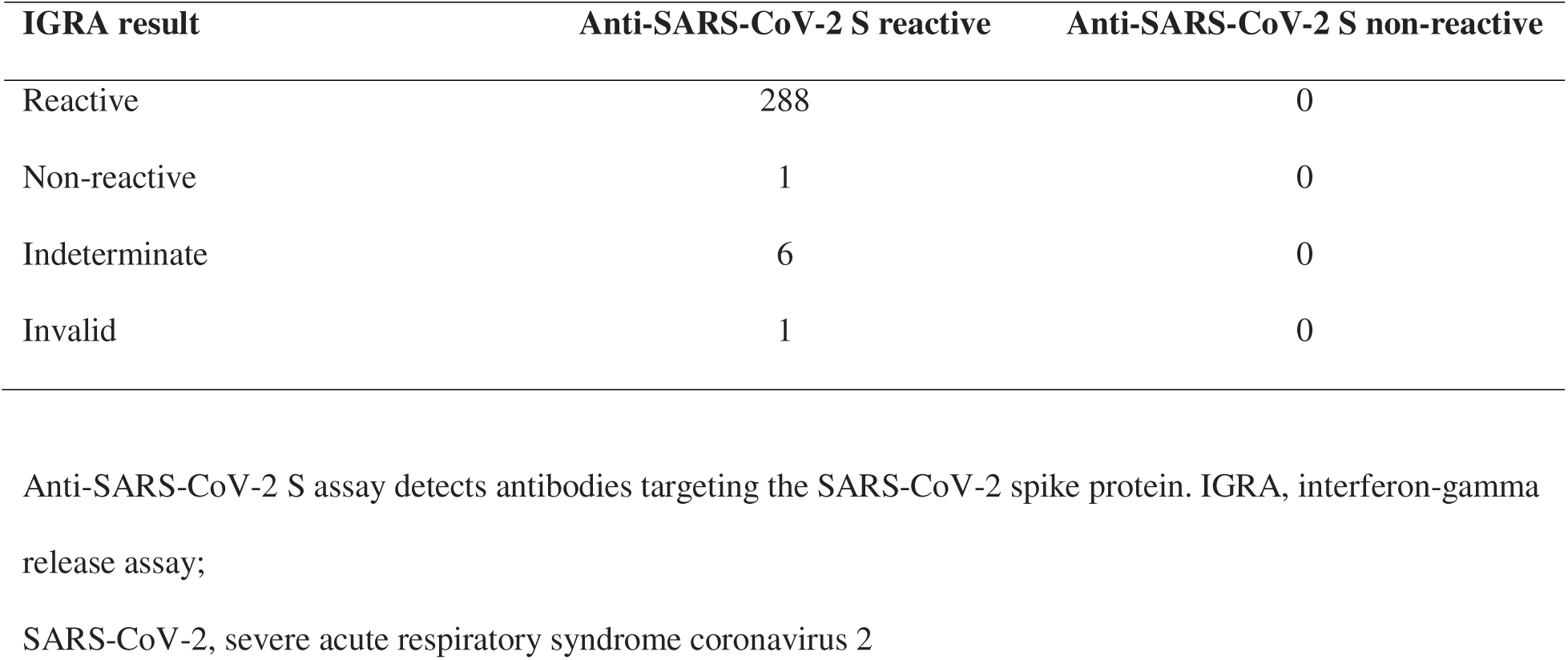
IGRA test results compared to Anti-SARS-CoV-2 S assay results in Cohort 2 (vaccines). Anti-SARS-CoV-2 assay detects antibodies targeting the SARS-CoV-2 spike protein. IGRA, interferon-gamma release assay; SARS-CoV-2, severe acute respiratory syndrome coronavirus 2.

**Fig. 3.**
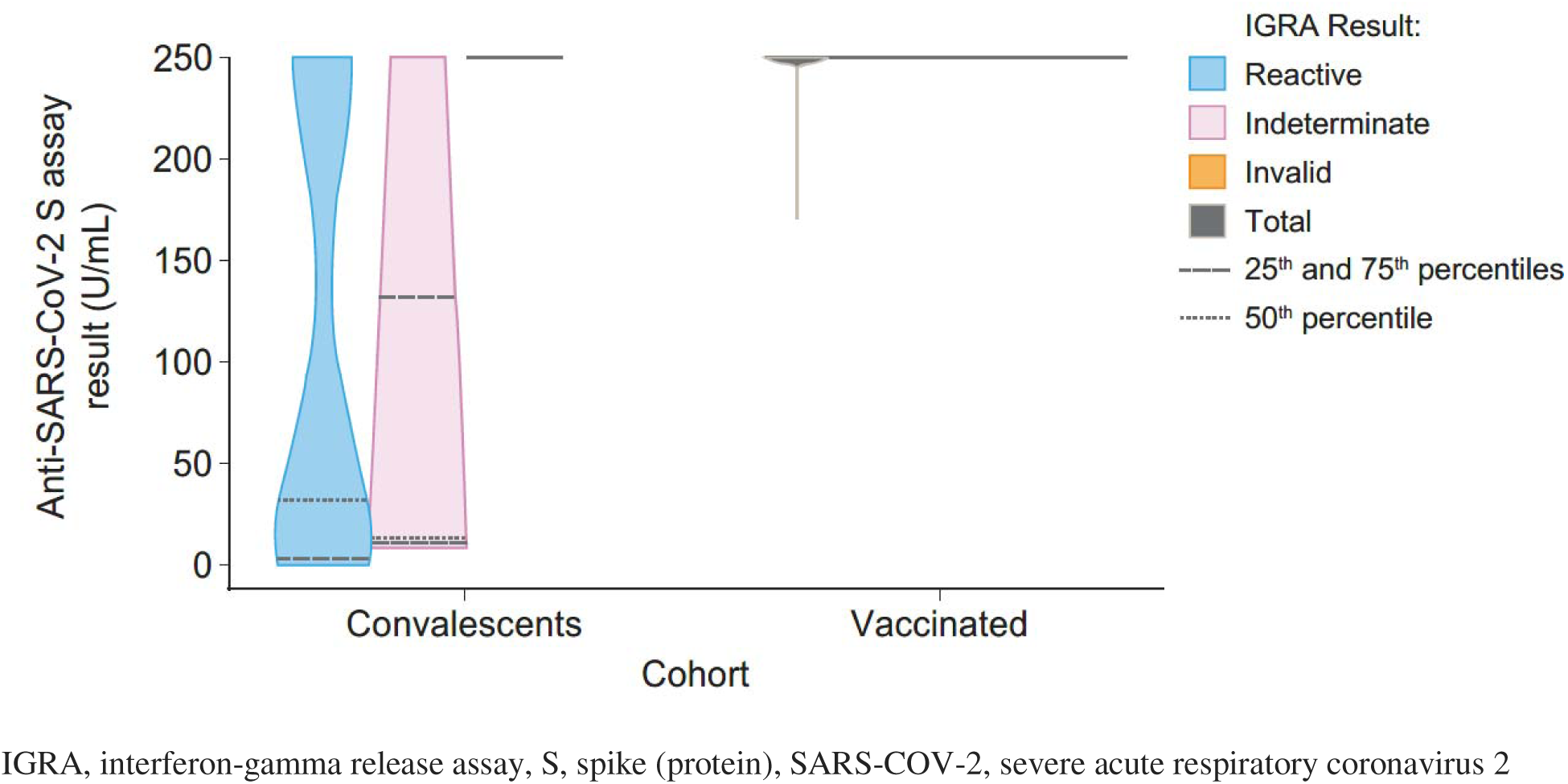
Distribution of Elecsys IGRA SARS-CoV-2 assay results compared to Elecsys Anti-SARS-CoV-2 S assay results. IGRA, interferon-gamma release assay, S, spike (protein), SARS-COV-2, severe acute respiratory coronavirus 2

### Specificity

In total, 78 whole blood samples were tested, and 77 valid results were obtained. Of these, 71 were non-reactive, 3 were reactive, and 3 were indeterminate. The clinical specificity was 92.21% (95% CI: 83.81 – 97.09) as also shown in **Table 4**.

**Table 4.** Clinical specificity of the IGRA SARS-CoV-2 assay. IGRA, interferon-gamma release assay; SARS-CoV-2, severe acute respiratory syndrome coronavirus 2.

| Cohort | N | Reactive | Non-reactive | Indeterminate<sup>a</sup> | Invalid<sup>b</sup> | Clinical sensitivity,<br>% (95% CI)<sup>c</sup> |
| --- | --- | --- | --- | --- | --- | --- |
| Negative donors | 78 | 3 | 71 | 3 | 1 | 92.21 (83.81–97.09) |

<sup>a</sup>High baseline IFN-γ in negative control
<sup>b</sup>Positive control below threshold, indicative of insufficient number of fit T cells
<sup>c</sup>Invalid samples were excluded from the sensitivity calculations
95% CI determined using Clopper–Pearson exact method
CI, confidence interval; IFN-γ , interferon-gamma; IGRA, interferon-gamma release assay; SARS-CoV-2, severe acute respiratory syndrome coronavirus 2

## Discussion

The Elecsys IGRA SARS-CoV-2 assay has both high sensitivity and high specificity in detecting SARS-CoV-2– elicited T-cell response, although there remains a need to validate the assay results in larger cohorts, including immunocompromised patient cohorts. Reactive results on the Elecsys IGRA SARS-CoV-2 assay were maintained up to 307 days post last dose vaccination and up to 736 days post-symptom onset. Measuring T-cell responses in convalescent and vaccinated individuals is feasible and can, in conjunction with serology assays, be meaningful in providing a more complete picture of the immune status after natural infections or vaccinations.

T-cell responses appear early in the course of infection, and their presence is correlated with protection; T-cell responses are weaker in severe disease occurring with lymphopenia. Further, the wide range of viral proteins recognized by T cells can provide protection against severe disease from viral variants, since T-cell memory is well sustained and, was estimated to recognize around 30 epitopes within each individual in one study [53]. Vaccine efficacy is derived from the adaptive immune response, which also determines the clinical outcome after SARS-CoV-2 infection. Currently available COVID-19 vaccines elicit robust T-cell responses that likely contribute to protection against hospitalization or death, and novel or heterologous regimens offer the potential to further enhance cellular responses [53].

Among indicators of the T-cell response to SARS-CoV-2, IFN-γ is a diverse cytokine that directs a wide array of cellular activities across the cell cycle, including immunomodulation and coordination of the innate and adaptive immune responses [49]. In addition to the promising results shown by the Elecsys IGRA SARS-CoV-2 assay for the detection of IFN-γ, a different IGRA T-cell assay recently demonstrated 100% specificity with high sensitivity to infection less than 5 months pre-test [47]. Another similar assay showed results that indicated SARS-CoV-2 seropositivity is correlated with broad T-cell reactivity for more than 200 days after infection [54]. However, T-cell assays are diverse, particularly functional T-cell assays. The performance and applicability for specific clinical use cases, depends on the nature of the assay stimulation design. Additionally, differences in human leukocyte antigen subtype prevalence indicate the need for test validation in diverse ethnic groups. Efforts are ongoing to validate our findings in a larger data set. In addition, these results call for future studies to demonstrate assay robustness in other ethnic groups and patient populations, particularly those with various mechanisms of immunodeficiencies or immune-dysregulations and the pediatric population.

Carretero et al. recently demonstrated that the Elecsys IGRA SARS-CoV-2 assay returned the highest number of SARS-CoV-2–positive test results (151/179; 84.3%) among different T-cell assays in vaccinated individuals, with varying levels of immunocompromised conditions. The authors concluded that the Elecsys IGRA SARS-CoV-2 assay may be particularly suited to revealing low-magnitude T-cell responses that may be present in immunosuppressed individuals, such as hematological patients. Further analysis using other methods revealed that the frequencies of SARS-CoV-2–reactive IFN-γ T cells and IFN-γ concentrations correlated moderately for CD4+ T cells but corelated weakly for CD8+ T cells [45]. Favresse et al. noted that in 54 participants of the CRO-VAX-HCP study who received the second and bivalent-adapted BNT162b2 booster, individuals with no history of SARS-CoV-2 infection presented with a significant waning of the humoral and cellular response after 6 months and were more likely to benefit from a second booster in terms of cellular immunity [55]. These data can help competent national authorities in their recommendation regarding the administration of an additional booster [51, 55].

## Conclusion

A more complete picture of T-cell response potential obtained through the Elecsys IGRA SARS-CoV-2 assay can aid physicians in making better-informed decisions for COVID-19 patient management in conjunction with other clinical information and additional testing results. Immediate clinical opportunities for IGRA assays include vaccination counseling of patients with immune disorders and guidance of treatment decisions and regular clinical care in patients with antibody deficiencies.

## Acknowledgments

Medical writing support was provided by Sumedha Sinha under the direction of the authors, and editorial support from Holly McAlister of inScience Communications, London and was funded by Roche Diagnostics International.

## Author Contributions

ME, QF, and US-S contributed to the study conception and design. All authors contributed to material preparation, data collection and analysis. All authors contributed to the preparation of the manuscript, and all authors read and approved the final manuscript.

## Ethics approval

The ethics committee Bayrische Landesärztekammer (Bavarian State Medical Association) gave ethical approval for this study.

## Informed Consent

All procedures were performed in accordance with the relevant guidelines and regulations, and each participant provided informed consent.

## Data availability

The study was conducted in accordance with applicable regulations. For more information on the study and data sharing, qualified researchers may contact the corresponding author, Dr Qian Feng.

